# MOE-ECG: Multi-Objective Ensemble Fusion for Robust Atrial Fibrillation Detection Using Electrocardiograms

**DOI:** 10.64898/2026.03.28.26349522

**Authors:** Abdolrahman Peimankar, Naser Hossein Motlagh, Smith K. Khare, Nicolai Spicher, Helena Dominguez, Vahid Abolghasemi, Koichi Fujiwara, Daniel Teichmann, Rahim Rahmani, Sadasivan Puthusserypady

## Abstract

**Background:** Atrial fibrillation (AFib) is the most common sustained arrhythmia in the world, imposing a heavy clinical and economic burden on global healthcare systems. Early detection of AFib can reduce mortality and morbidity, while helping to alleviate the growing economic burden of cardiovascular diseases. With the increasing availability of digital health technologies, computational solutions have great potential to support the timely diagnosis of cardiac abnormalities.

**Objectives:** With the increasing availability of electrocardiogram (ECG) data from clinical and wearable devices, manual interpretation has become impractical due to its time-consuming and subjective nature. Existing automated approaches often rely on single classifiers or fixed ensembles that primarily optimize predictive accuracy while neglecting model diversity, which leads to limited robustness and generalization across heterogeneous datasets. Therefore, this study aims to develop a robust and diversity-aware framework for automatic AFib detection that simultaneously improves classification performance and model generalizability. To this end, we propose MOE-ECG, a multi-objective ensemble selection and fusion framework that explicitly optimizes both predictive performance and inter-model diversity for reliable AFib detection from ECG recordings.

**Methods:** The proposed multi-objective ensemble (MOE) framework uses ensemble selection as a bi-objective optimization problem and employs multi-objective particle swarm optimization to identify complementary classifiers from a heterogeneous model pool. Unlike conventional ensembles, it explicitly optimizes both predictive performance and diversity and integrates Dempster–Shafer theory for uncertainty-aware decision fusion. After filtering the ECG signals to remove baseline wander and noise, they were segmented into windows of 20, 60, and 120 heartbeats with 50% overlap. The proposed approach was evaluated over five independent runs to assess its stability and generalization. Fifteen statistical and nonlinear features were obtained from the RR-intervals of the pre-processed ECG signals, of which eight features were selected with correlation analysis to capture subtle information from the ECG data. We trained and evaluated the performance of the proposed model in three open source databases, namely, the MIT-BIH Atrial Fibrillation Database, Saitama Heart Database Atrial Fibrillation, and Long-Term AF Database.

**Results:** The proposed approach achieved the best overall performance on 60-beat segments, with an average accuracy of 89.85%, precision of 91.14%, recall of 94.19%, an *F*_1_-score of 92.64%, and area under the curve (AUC) of around 0.95. Statistical analysis using Holm-adjusted Wilcoxon tests confirmed significant improvements (*p <* 0.05) compared to both the best individual classifier and the unoptimized average ensemble of all classifiers. These findings show that the proposed selection and evaluation methodology, rather than group aggregation alone, is the key driver of performance improvements.

**Conclusion:** The results obtained demonstrate that the MOE-ECG model offers a robust, accurate, and reliable solution for the detection of AFib from short ECG segments. The empirical findings, in general, confirm that multi-objective ensemble fusion enhances diagnostic performance and offers robust predictions that will open up possibilities for real-time AFib detection in clinical and tele-health settings.

## 1. Introduction

Atrial fibrillation (AFib) is the most common sustained cardiac arrhythmia and represents a major global health burden, which affects more than 33 million people worldwide [1]. The prevalence increases with age and comorbidities such as hypertension, diabetes, and heart failure, and continues to be one of the leading causes of stroke and heart-related morbidity and mortality [2, 3]. Its intermittent and often asymptomatic nature poses a significant challenge to diagnosis, as many patients remain undiagnosed until serious complications occur. Therefore, early and reliable detection of AFib can reduce adverse clinical outcomes and substantially alleviate healthcare costs.

Traditionally, AFib diagnosis has relied on visual inspection of electrocardiograms (ECG) by clinicians to identify patterns such as irregular RR-intervals and absence of P waves [4, 5]. However, the generation of large amounts of ECG data due to continuous monitoring, which is needed for the detection of paroxysmal AFib, makes a manual review impractical. In addition to traditional 10s 12-lead ECG measurements, Holter monitors and cardiac patches that continuously measure ECG over multiple days, as well as handheld devices and smart watches for point measurements, found their way into clinical guidelines and practice [6]. This has motivated the development of automated and intelligent systems capable of accurate and efficient detection of AFib from ECG recordings [7, 8]. Over the past two decades, a diverse array of signal processing and machine learning (ML) frameworks has emerged to automate AFib detection. These methodologies utilize a combination of linear and nonlinear features extracted across the time and frequency domains, derived from both RR-interval sequences and raw ECG signals [9, 10, 11].

While common ML algorithms such as support vector machines (SVM), random forests (RF), and boosting ensembles have shown promising results when combined with carefully engineered features [12, 13], most existing approaches either rely on single classifiers or employ fixed or homogeneous ensembles optimized for a single objective (typically classification accuracy). In many cases, ensemble construction is heuristic or based on simple averaging, without explicitly modeling the trade-off between predictive performance and classifier diversity, which is crucial for achieving robustness and reproducibility in heterogeneous ECG datasets [14].

Recent research indicates that ensemble diversity is fundamental to enhancing classification stability, mitigating overfitting, and improving the sensitivity of the model toward rare or irregular patterns [14, 15, 16]. Multi-objective optimization approaches, such as the Multi-Objective Particle Swarm Optimization (MOPSO) algorithm, are efficient methods to explore trade-offs between conflicting goals, for example, maximizing accuracy while maintaining high classifier diversity [17, 18]. In the medical domain, MOPSO-based frameworks have been successfully applied to feature selection and diagnosis tasks on heterogeneous biomedical datasets [19, 20, 21]. More recently, MOPSO has also been used to design or select ensembles in high-dimensional classification problems [22, 23]. However, its utility in constructing diversity-aware ECG ensembles for the detection of AFib mostly remains unexplored [24].

In this study, we propose MOE-ECG, a novel Multi-Objective Ensemble Fusion framework for AFib detection. The main contributions of this work are as follows: (1) We formulate ensemble selection as a bi-objective optimization problem that explicitly balances predictive performance and classifier diversity, addressing a key limitation of conventional accuracy-driven or heuristic ensemble methods. (2) We introduce a diversity-aware ensemble selection strategy based on MOPSO, which automatically identifies a compact subset of complementary classifiers from a heterogeneous pool of 22 models, avoiding predefined or fixed ensemble structures. (3) We integrate Dempster–Shafer evidence theory for uncertainty-aware fusion of classifier outputs, enabling more robust probabilistic decision-making. (4) We demonstrate the effectiveness and generalizability of the proposed framework through systematic cross-dataset evaluation, achieving improved performance compared to both the single best classifier and standard averaging ensembles.

The remainder of this paper is organized as follows: Section 2 provides the methodology of the proposed AFib detection framework in addition to the detailed description of the datasets, features, and classifiers used in this work. The results of the proposed model are presented and discussed in Section 3, followed by a conclusion in Section 4.

## 2. Materials and methods

### 2.1. Dataset

In this study, three publicly available ECG databases from PhysioNet [25] were used to evaluate the proposed MOE-ECG framework. The MIT-BIH Atrial Fibrillation Database (AFDB) served as the training dataset. AFDB contains 25 long-term two-channel ECG recordings of approximately 10 hours each, sampled at 250 Hz, with beat-by-beat annotations indicating AFib and non-AFib rhythms [26]. Two recordings were omitted from the analysis due to a lack of corresponding annotation files. The Saitama Heart Database Atrial Fibrillation (SHDB-AF) was used for model validation [27, 28]. This dataset comprises 128 Holter two-channel ECG recordings collected from 122 unique patients, each with a duration of exactly 24 hours sampled at 125 Hz. Expert-labeled AFib/Atrial Flutter annotations are available for 98 of the 128 recordings. Independent generalization performance was assessed using the Long-Term Atrial Fibrillation Database (LTAFDB), which includes 84 two-channel 24-hour Holter ECG recordings sampled at 128 Hz [29]. Each record contains detailed beat annotations enabling precise identification of AFib and normal sinus rhythms. All three datasets provide reference annotations indicating the timing and type of each heartbeat. A summary of the datasets used in this study is presented in Table 1.

**Table 1:** Summary of the three ECG datasets used in this study.

| Dataset | Record length (hrs) | Subjects # | Sampling rate (Hz) | AFib rhythms | AFib length (hrs) |
| --- | --- | --- | --- | --- | --- |
| AFDB | ~ 10 | 23 | 250 | 291 | ~ 39 |
| SHDB-AF | ~ 24 | 122 | 125 | 809 | ~ 1459 |
| LTAfDB | ~ 24 | 84 | 128 | 7339 | ~ 1031 |

### 2.2. Data preprocessing

RR-interval (RRI) sequences were derived from the raw ECG signals to serve as the foundational data for all subsequent feature extraction. R-peak positions were obtained using the PhysioNet WFDB Toolbox [30], which implements the classical Pan–Tompkins algorithm for QRS detection [25, 31]. For recordings with available reference annotations, the ground-truth R-peak locations provided by experts were utilized directly. To maintain consistency across all datasets, only a single ECG lead was utilised from each record. The Modified Limb Lead II (MLII) was adopted for AFDB since it is the standard monitoring lead in PhysioNet arrhythmia studies and provides high R-wave amplitude for reliable R-peak detection. The CC5 lead was used for SHDB-AF due to its strong ventricular morphology and favorable signal-to-noise characteristics in Holter monitoring. For LTAFDB, the first Holter channel was selected as the primary acquisition channel to ensure reproducibility across subjects. All ECG recordings were resampled to a common sampling frequency of 250 Hz to ensure uniformity of R-peak timing across datasets. RRIs were computed as the time differences between consecutive R-peaks. Each ECG recording was segmented into overlapping windows of 20, 60, and 120 consecutive RRIs to capture rhythm dynamics at multiple temporal scales. These window lengths approximately correspond to short (∼15–20 s), medium (∼45–60 s), and longer (∼90–120 s) cardiac contexts at typical heart rates, which enables a trade-off between rapid AF detection and statistically stable variability estimation. Multi-scale segmentation is commonly adopted in RR/HRV-based arrhythmia analysis to balance responsiveness and robustness [32, 33, 34]. A segment was labeled as AFib if more than 50% of its constituent RRIs were annotated as AFib; otherwise, labeled as Normal. This majority-based labeling approach is commonly adopted in AFib detection studies and ensures reliable window-level rhythm annotation. For each segment length, the number of segments belonging to Normal and AFib classes was computed for the three datasets. These statistics are summarised in Table 2. As expected, AFDB exhibited moderate class imbalance, while SHDB-AF and LTAFDB yielded a more balanced distribution of AFib and Normal segments due to their longer continuous Holter recordings. Example RRI patterns for Normal sinus rhythm (NSR) and AFib rhythms are shown in Figure 1, where the first exhibits regular and stable RR-intervals, whereas the latter displays beat-to-beat irregularity.

**Table 2:** Number of rhythms per class for different segment lengths among the four datasets.

| Dataset | $RRI=20$ | | $RRI=60$ | | $RRI=120$ | |
| --- | --- | --- | --- | --- | --- | --- |
|  | Normal | AFib | Normal | AFib | Normal | AFib |
| AFDB | 45432 (64.1%) | 25424 (35.9%) | 22739 (64.2%) | 12689 (35.8%) | 7578 (64.2%) | 4231 (35.8%) |
| SHDB-AF | 382004 (75.7%) | 122737 (24.3%) | 127319 (75.7%) | 40928 (24.3%) | 63653 (75.7%) | 20470 (24.3%) |
| LTAfDB | 291375 (46.7%) | 332720 (53.3%) | 145844 (46.7%) | 166203 (53.3%) | 48667 (46.8%) | 55348 (53.2%) |

**Figure 1:**
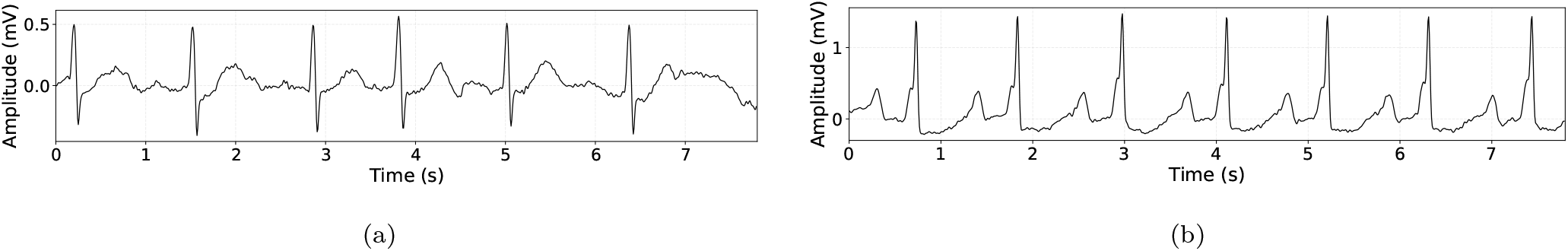
An example of (a) AFib and (b) NSR segments. These are excerpts from the AFDB dataset. The y-axis represents the ECG amplitude in mV and the x-axis shows the time in seconds.

### 2.3. Feature extraction and selection

A total of fifteen RRI-based features were extracted from each segment to capture the statistical, temporal, and nonlinear characteristics of heart rate dynamics. These features include standard statistical measures (e.g., Mean_RR, Min, Max) and time-domain variability descriptors (e.g., SD_RR, RMSSD, nRMSSD, NN50, pNN50), which are widely used in heart rate variability (HRV) analysis [35, 36]. In addition, distribution-based measures (MeanAD, MedianAD) [37] and nonlinear and complexity-related features such as entropy, CVI, CVS, and the SD2/SD1 ratio derived from Poincaré plots [38, 39, 40] were included. The average variation rate (AVRR) was also considered to capture rapid beat-to-beat fluctuations characteristic of AFib [41].

Together, these features provide a compact yet informative representation of rhythm irregularity, variability, and complexity, which are key discriminative markers between NSR and AFib. A detailed summary of all extracted features, including their definitions and mathematical formulations, is provided in Table 3.

**Table 3:**
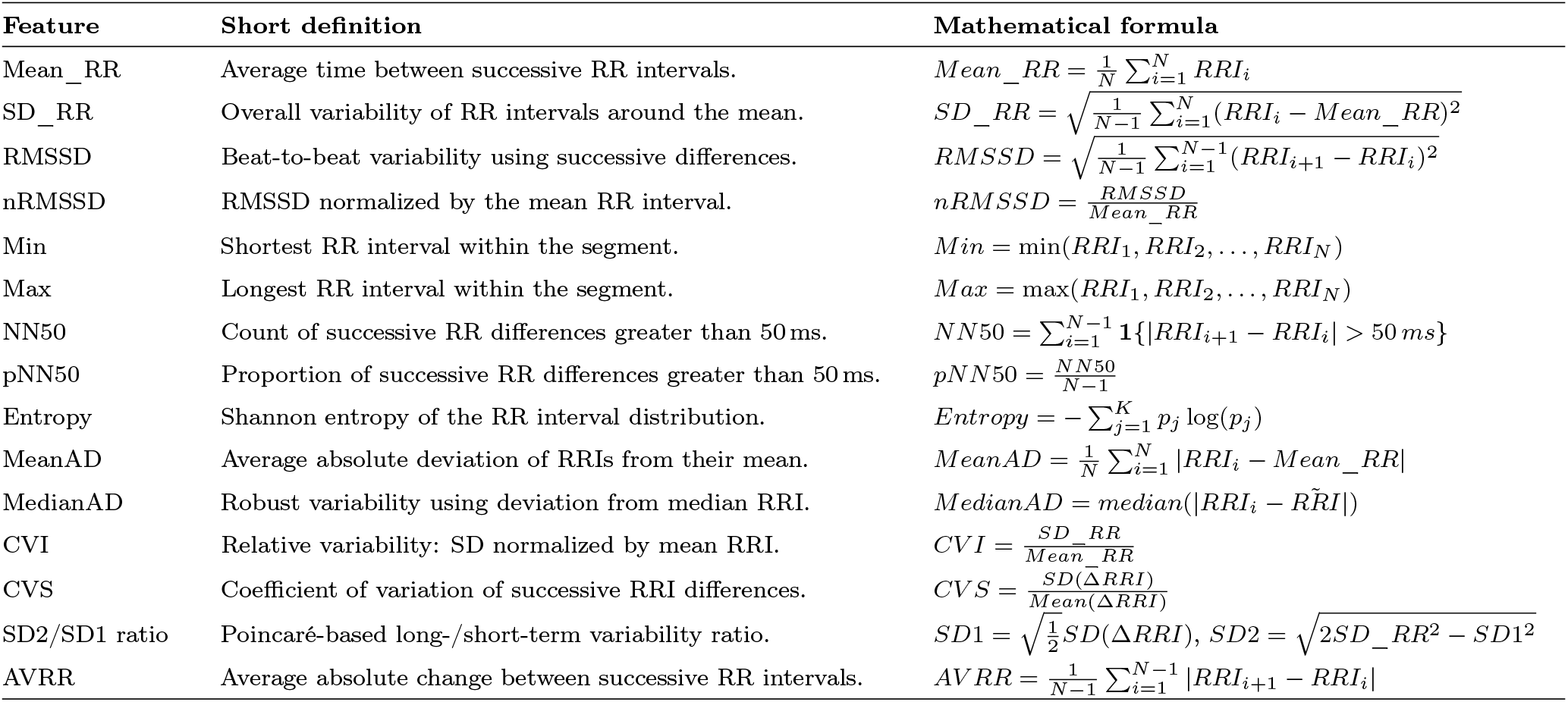
Summary of the extracted RRI-based features used for AFib detection.

Figure 2 shows the Pearson pairwise correlations among all extracted features, along with scatter plots and marginal histograms. The slope of the fitted lines represents the correlation coefficients, while asterisks denote statistical significance, with larger values and more asterisks indicating stronger relationships. The figure also highlights both the relationship of individual features with the *Class* label (NSR vs. AFib) and the degree of inter-feature redundancy.

**Figure 2:**
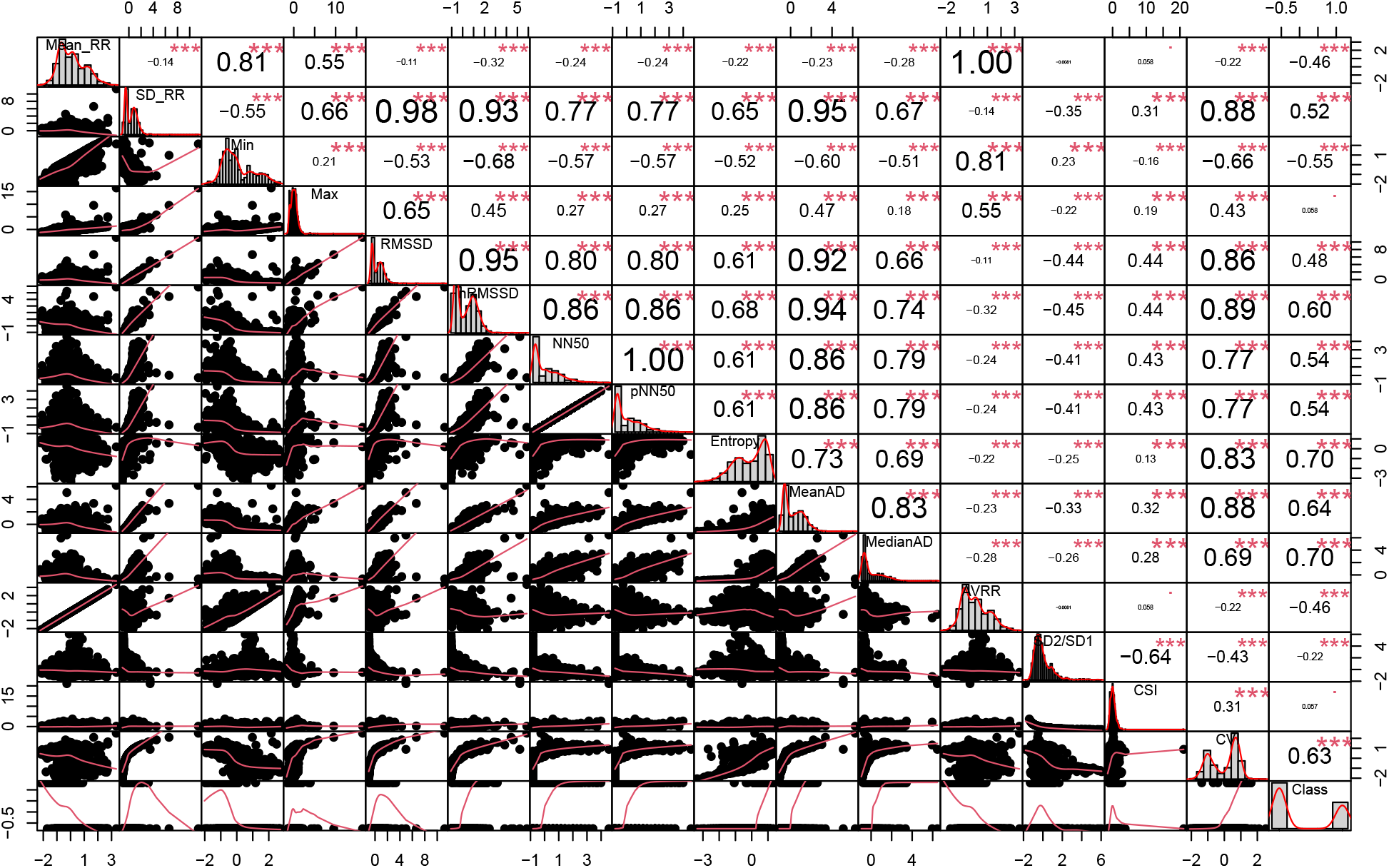
The Pearson pairwise correlation coefficients are displayed together with the scatter plots of each feature pair and the marginal histograms along the diagonal. The slope of the least-squares fitted line in each scatter plot equals the corresponding correlation coefficient. Larger absolute values and a greater number of asterisks indicate more statistically significant correlations between feature pairs. This visualization highlights strongly correlated feature clusters, which were subsequently pruned to reduce redundancy during feature selection.

Several strongly correlated clusters are observed, particularly among short-term variability features (RMSSD, nRMSSD, NN50, pNN50, MeanAD, and MedianAD), all showing high mutual correlations (|*r*| *>* 0.85). Similarly, SD_RR and Min form a highly collinear pair, while Entropy and CVI exhibit substantial shared information.

To reduce redundancy, a two-stage feature selection strategy was applied. First, features were ranked based on their absolute Pearson correlation with the *Class* label. Second, redundancy pruning removed features with |*r*| *>* 0.85 relative to higher-ranked features. This process yielded a compact subset of ten features (MedianAD, Entropy, CVI, MeanAD, nRMSSD, NN50, RMSSD, SD_RR, Mean_RR, and AVRR), capturing key temporal, statistical, and nonlinear characteristics of AFib while minimizing multicollinearity and preserving interpretability.

Figure 3 presents log-scaled boxplots of the selected ten RRI-based features for NSR and AFib segments. Several features (e.g., MedianAD, MeanAD, RMSSD, nRMSSD, NN50, and AVRR) show higher values in AFib, reflecting increased beat-to-beat irregularity. Entropy and CVI exhibit greater dispersion in AFib, indicating higher complexity, while Mean_RR shows a slight decrease, consistent with faster ventricular response. Overall, the distributions demonstrate that the selected features capture complementary and discriminative AFib-related characteristics.

**Figure 3:**
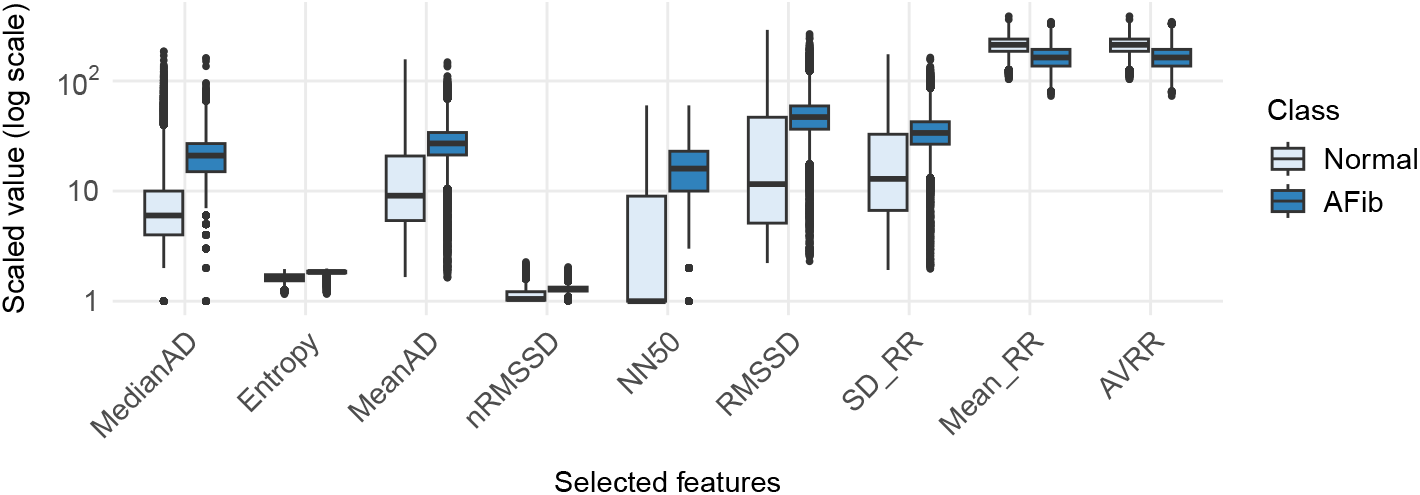
Log-scaled boxplots of the selected ten RRI-based features for Normal and AFib segments. Clear separation between the two groups is visible for several features, particularly MedianAD, MeanAD, RMSSD, nRMSSD, NN50, and AVRR, which all show higher values in AFib due to increased beat-to-beat irregularity. Nonlinear measures such as Entropy and CVI also exhibit elevated values and wider dispersion in AFib, reflecting the greater complexity of arrhythmic RRI dynamics. Mean_RR shows a modest decrease for AFib, consistent with faster ventricular response. Overall, the distributions demonstrate that the selected features capture distinct and complementary AF-related characteristics suitable for classification.

To further evaluate discriminative power, low-dimensional embeddings using UMAP [42] and t-SNE [43] are shown in Figures 4a and 4b. Both visualizations reveal a clear tendency for AFib and NSR segments to form separable structures, with AFib samples appearing more dispersed due to their irregular dynamics. Despite some overlap, these embeddings confirm that the selected features capture meaningful structure for distinguishing between the two classes.

**Figure 4:**
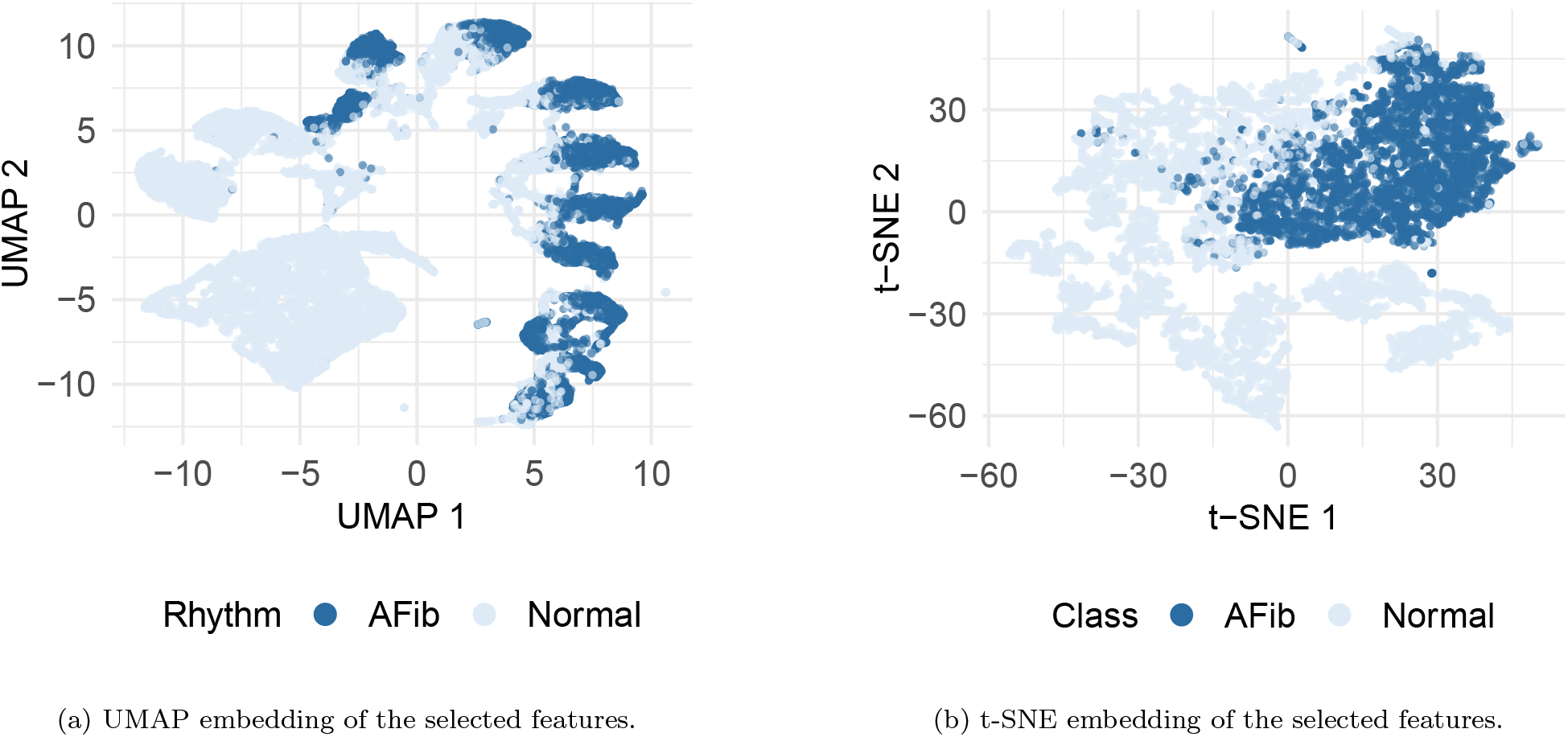
Low-dimensional visualization of the ten selected RRI-based features. (a) UMAP and (b) t-SNE both show that AFib and Normal segments form distinguishable patterns, with AFib samples exhibiting more dispersed and heterogeneous structure due to their irregular RRI dynamics. These visualizations demonstrate that the selected features capture meaningful discriminative information suitable for AFib detection.

### 2.4. Multi-objective ensemble classifier selection

To design a robust and accurate AFib detection system, we employed a multi-objective ensemble learning strategy based on a heterogeneous set of 22 classifiers and optimized them simultaneously for predictive performance and classifier diversity. AFib produces nonlinear and irregular fluctuations in the RRI series; therefore, using a diverse model pool helps capture complementary patterns that a single classifier family cannot learn effectively. This motivation is well-supported by ensemble learning theory [14, 44, 45].

A comprehensive ensemble of twenty-two classifiers was evaluated, comprising linear, probabilistic, and distance-based models, as well as tree-based architectures—including advanced gradient boosting frameworks such as XGBoost and LightGBM. Furthermore, the selection included Tab-Net [46] and four deep learning architectures: MLP, DeepNet, VGG, and ResNet. To optimize performance, hyperparameter tuning was conducted via grid search [47, 48], utilizing a stratified 5-fold cross-validation scheme to ensure robust evaluation. The full set of hyperparameter ranges is summarized in Table 4. The best-performing configuration based on validation F_1_-score was preserved for the ensemble selection stage.

**Table 4:** Hyperparameter ranges explored for each classifier during grid search.

| Classifier | Hyperparameters |
| --- | --- |
| MLP | Number of hidden layers = [1,2,3]; Number of units = [32,64,128]; Activation function = [relu,elu]; Optimizer = [adam,rmsprop]; Batch size = [32,64]; Number of epochs = [10,20]. |
| Deep Neural Network | Number of layers = [2,3,4]; Number of units = [128,256]; Activation function = [relu,elu]; Optimizer = [adam,rmsprop]; Batch size = [32,64]; Number of epochs = [10,20]. |
| VGG | Convolution block sizes = [[32,64],[64,128]]; Dense units = [256,512]; Dropout rate = [0.3,0.4]; Activation function = [relu]; Optimizer = [adam]; Batch size = [32]; Number of epochs = [10]. |
| ResNet | Number of filters = [32,64]; Number of residual blocks = [2,3]; Dense units = [64,128]; Dropout rate = [0.2,0.3]; Activation function = [relu]; Optimizer = [adam]; Batch size = [32]; Number of epochs = [10]. |
| TabNet | Feature dimension ($n_d$) = [8,16,32]; Attention dimension ($n_a$) = [8,16,32]; Number of decision steps = [3,5]; Gamma (relaxation) = [1.0,1.5]; Sparsity regularization = [1e-3,1e-4]; Momentum = [0.02]; Mask type = [sparsemax]. |
| Logistic Regression | Regularization strength $C$ = [0.001,0.01,0.1,1]; Penalty type = [l1,l2]; Solver = [liblinear,saga]; Maximum number of iterations = [200,500]. |
| Random Forest | Number of estimators = [100,200,300]; Maximum depth = [None,10,20,30]; Minimum samples split = [2,5,10]; Minimum samples per leaf = [1,2,4]; Maximum number of features = [sqrt,log2]. |
| XGBoost | Number of estimators = [100,200]; Maximum depth = [3,6,9]; Learning rate = [0.01,0.1,0.3]; Subsample ratio = [0.8,1.0]; Column subsample (per tree) = [0.8,1.0]; Gamma (tree regularization) = [0,0.1,0.2]. |
| LightGBM | Number of estimators = [100,200]; Number of leaves = [31,63,127]; Learning rate = [0.01,0.1]; Subsample ratio = [0.8,1.0]; Feature subsample ratio = [0.8,1.0]; Minimum child samples = [20,50]. |
| Decision Tree | Maximum depth = [None,10,20,30]; Minimum samples split = [2,5,10]; Minimum samples per leaf = [1,2,4]; Splitting criterion = [gini,entropy]. |
| K-Nearest Neighbors | Number of neighbors = [3,5,7,9]; Weighting scheme = [uniform,distance]; Distance metric = [euclidean,manhattan]; Minkowski power $p$ = [1,2]. |
| GaussianNB | Variance smoothing = [1e-9]. |
| BernoulliNB | Smoothing parameter $\alpha$ = [0.1,0.5,1.0]; Binarization threshold = [0.0,0.5]. |
| LinearSVC | Regularization strength $C$ = [0.1,1,10]; Loss function = [hinge,squared_hinge]; Maximum number of iterations = [1000,2000]. |
| RidgeClassifier | Regularization strength $\alpha$ = [0.1,1.0,10.0]; Solver = [auto,cholesky]; Maximum number of iterations = [None,500]. |
| SGDClassifier | Loss function = [hinge,log_loss,modified_huber]; Penalty = [l2,l1,elasticnet]; Regularization parameter $\alpha$ = [1e-4,1e-3,1e-2]; Maximum number of iterations = [1000]; Tolerance = [1e-3]. |
| ExtraTrees | Number of estimators = [100,200]; Maximum depth = [None,10,20]; Minimum samples split = [2,5]; Minimum samples per leaf = [1,2]; Maximum number of features = [sqrt,log2]. |
| AdaBoost | Number of estimators = [50,100]; Learning rate = [0.1,1.0]; Boosting algorithm = [SAMME,SAMME.R]. |
| Gradient Boosting | Number of estimators = [100,200]; Learning rate = [0.01,0.1]; Maximum depth = [3,5]; Minimum samples split = [2,5]; Minimum samples per leaf = [1,2]. |
| Bagging | Number of estimators = [10,20,50]; Maximum samples ratio = [0.5,0.7,1.0]; Maximum features ratio = [0.5,0.7,1.0]. |
| LDA | Solver = [svd,lsqr,eigen]; Shrinkage = [None,auto]; Tolerance = [1e-4,1e-3]. |
| QDA | Regularization parameter = [0,0.1,0.2]; Tolerance = [1e-4,1e-3]. |

After hyperparameter optimization, classifier selection was formulated as a bi-objective optimization problem. The selected ensemble should both maximize classification performance and maximize classifier diversity.

Let **s** = [*s*_1_, *s*_2_, …, *s*_*L*_] be the binary classifier selection vector, where *s*_*i*_ = 1 indicates that the *i*th classifier is included in the ensemble and *s*_*i*_ = 0 otherwise, and *F*_1_ is the computed *F*_1_-score of the ensemble composed of the selected classifiers. The ensemble performance objective is defined as the maximization of the validation *F*_1_-score:

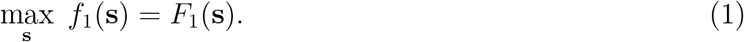

There are two different approaches to measure the diversity of the selected classifiers; pairwise and non-pairwise [14]. In this paper, a pairwise disagreement among classifiers is computed [49]. For selected classifiers in **s**, let *P* ^(*i*)^ ∈ {0, 1}^*N*^ denote the hard prediction vector of classifier *i*, and let *L* =∑_*i*_ *s*_*i*_ be the total number of classifiers selected in the ensemble.

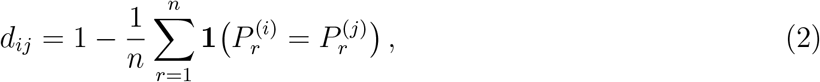

Then, the overall ensemble diversity (average pairwise disagreement) is:

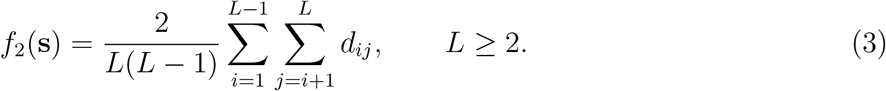

### 2.5. Pareto optimality in multi-objective optimization

Pareto optimality is a fundamental concept in multi-objective optimization and forms the foundation for identifying the optimal trade-offs between conflicting design goals [50, 51]. Within the scope of the proposed ensemble learning framework, performance and diversity are two such objectives that cannot be fully optimized at the same time without making some level of compromise. It is, therefore, essential to introduce the concept of Pareto optimality before detailing the MOPSO-based ensemble selection process.

Mathematically, a general multi-objective optimization problem with *P* objective functions and *N* inequality constraints can be formulated as follows [50, 52]:

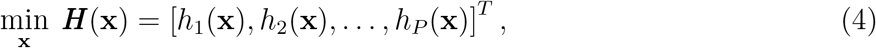

subject to

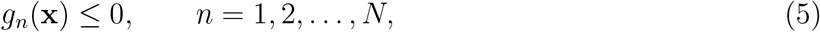

where ***x*** is an *m*-dimensional decision vector taken from a feasible region. The functions ***H***(***x***) represent the *P* conflicting objectives, while *g*_*n*_(***x***) are constraint functions. In multi-objective optimization, the objective functions may be linear or nonlinear, convex or non-convex.

In almost all real-world multi-objective optimization problems, including classifier ensemble selection, the objectives are mutually conflicting. As a result, one cannot optimize all objectives simultaneously. Instead, the goal is to identify solutions that offer the best possible trade-off. These solutions are referred to as *Pareto optimal* or *non-dominated* solutions.

To formally define these concepts, consider two objective vectors ***u*** = (*u*_1_, *u*_2_, …, *u*_*P*_) and ***v*** = (*v*_1_, *v*_2_, …, *v*_*P*_) associated with two feasible candidate solutions. For a minimization problem, the concept of dominance is defined as follows [50, 52]:

- *Dominance:* A vector ***u*** is said to dominate ***v*** (denoted ***u*** ≺ ***v***) if and only if ∀ *i* ∈ {1, 2, …, *L*}, *u*_*i*_ ≤ *v*_*i*_ ∧ ∃ *i* ∈ {1, 2, …, *L*}: *u*_*i*_ *< v*_*i*_.
- *Pareto optimality:* A candidate solution ***x***^⋆^ ∈ Θ is considered Pareto optimal if, within the feasible set Θ, there exists no alternative solution *x* such that the objective vector ***H***(***x***) = [*h*_1_(***x***), *h*_2_(***x***), …, *h*_*P*_ (***x***)] is strictly superior to ***H***(***x***^⋆^) = [*h*_1_(***x***^⋆^), *h*_2_(***x***^⋆^), …, *h*_*P*_ (***x***^⋆^)]. in all objectives and strictly better in at least one of them. In other words, ***x***^⋆^ is non-dominated with respect to all other feasible solutions.

The set of all non-dominated solutions forms the *Pareto front*. In the proposed ensemble learning framework, each point on the Pareto front represents a specific classifier subset characterized by a trade-off between ensemble performance and classifier diversity. Using MOPSO [51], the algorithm maintains an external repository of such non-dominated solutions, and the final ensemble is selected from this set based on validation F_1_-score. This ensures that the chosen ensemble is both accurate and diverse while adhering to the principles of multi-objective optimality.

### 2.6. Multi-objective PSO

To determine the best subset of the classifiers for the proposed ensemble, we use a multi-objective particle swarm optimization algorithm. MOPSO extends the classical PSO method, which is used to optimize a single objective. In contrast, MOPSO is designed to handle multiple, possibly conflicting objectives. In our case, the objectives correspond to the following: (i) maximizing predictive performance of the ensemble, and (ii) enhancing classifier diversity. Optimizing these two quantities jointly will ensure that the final ensemble is both accurate and composed of diverse models.

In the canonical PSO framework, each particle denotes a candidate solution in the search space, with its trajectory governed by iterative updates. The velocity at iteration *k* is determined by the weighted interplay of three distinct influences: (i) an inertia component that preserves momentum from previous iterations; (ii) a cognitive (or ‘nostalgia’) term that attracts the particle toward its personal historical optimum (*personal best*); and (iii) a social term that steers the swarm toward the overall global best (*global best*). Following the formulation by Kennedy and Eberhart [53], these update dynamics are defined as:

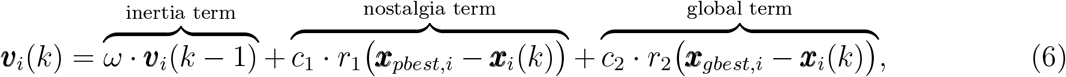

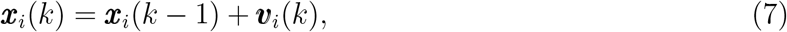

where *ω* is the inertia weight, **x**_*pbest,i*_ denotes the personal best position of particle *i*, **x**_*gbest,i*_ denotes the position of the swarm leader, and *r*_1_ and *r*_2_ are uniformly distributed random variables in [0, 1], and *c*_1_ and *c*_2_ represent the learning coefficients that determine the relative contribution of the cognitive (*personal best*) and social (*global best*) components. In this work, we employ parameter values consistent with established recommendations in earlier studies, which are *c*_1_ = *c*_2_ = 2 and *ω* = 0.8 [54].

Unlike classical PSO, the MOPSO algorithm does not rely on a single global best strategy. Instead, it maintains an external repository (or *archive*) of non-dominated solutions, forming an approximation of the Pareto-optimal set [55]. When selecting a leader for a particle, MOPSO samples from this repository rather than using a single best position since they are all non-dominated solutions. To improve the spread of solutions across the Pareto front, a region-based leader selection mechanism is adopted in which the objective space is divided into subregions [17, 51, 56]. Leaders from sparsely populated regions are given higher selection probability, which encourages search diversity and promotes the discovery of non-dominated solutions as *global best* candidates.

We also apply a mutation operator, as recommended in [17], to prevent premature convergence and to explore new areas of the search space. After each update, the dominated solutions are removed from the repository to ensure that only Pareto-efficient candidates remain. To prevent the repository from exceeding its predefined capacity, only the set of non-dominated leaders is retained, and the occupancy of the subregions is updated accordingly. Before executing the MOPSO algorithm, several key parameters must be specified. These include:

- *nPop*; population size
- *MaxIt*; maximum number of iterations
- *nRep*; repository (archive) size
- ***µ***; mutation rate
- ***β***; leader-selection pressure
- *nRegion*; number of subregions in the objective space
- ***c***_**1**_ and ***c***_**2**_; learning coefficients
- ***ω***; inertia weight

The overall MOPSO procedure used in this study are provide in Algorithm 1.

#### Algorithm 1

MOPSO algorithm

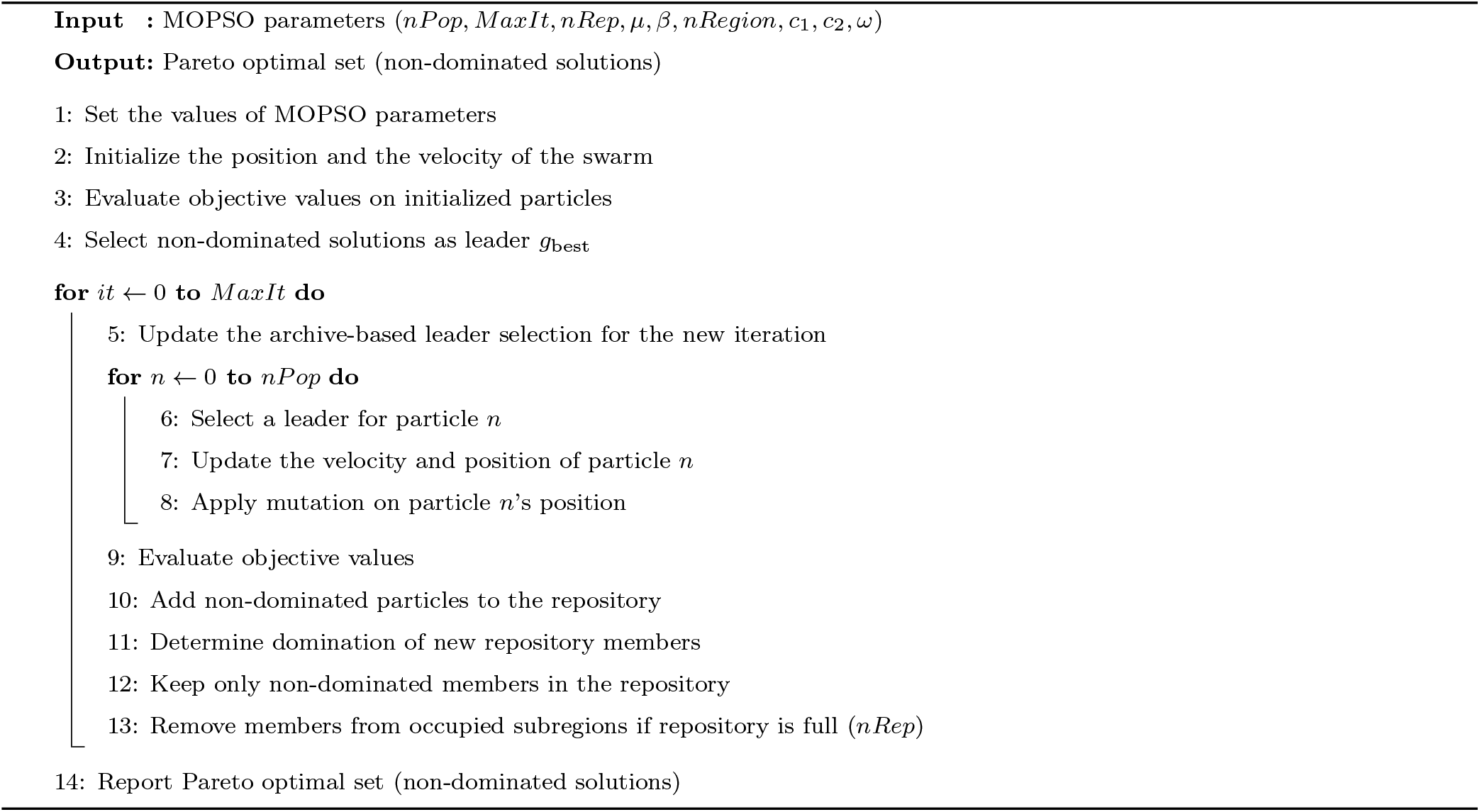

### 2.7. Dempster–Shafer combination rule

To fuse the outputs of the classifier ensemble selected by the MOPSO algorithm, we adopt the Dempster–Shafer Theory (DST) [57], which provides a rigorous framework for combining independent sources of evidence and quantifying uncertainty. Unlike traditional probabilistic fusion, DST allows each classifier to contribute a degree of belief to individual hypotheses as well as to sets of hypotheses, making it especially suitable for medical decision systems where uncertainty and disagreement between classifiers may occur.

DST is defined over a universal hypothesis set *X* = {Normal, AFib}. For any subset *A* ⊆ *X*, the mass function *m*: 2^*X*^ → [0, 1] assigns a basic probability value according to:

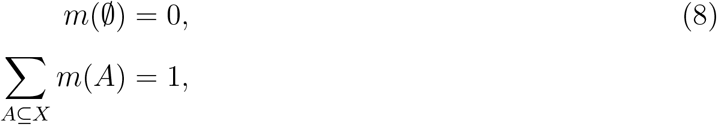

where *m*(*A*) represents the support committed exactly to subset *A*. In this study, the normalized confidence scores produced by each selected classifier are interpreted as basic mass assignments, allowing each classifier to contribute an independent piece of evidence regarding the rhythm class.

DST combines two independent mass functions *m*_1_ and *m*_2_ using the Dempster–Shafer combination rule. The combined mass assigned to a subset *A* ⊆ *X* is given by:

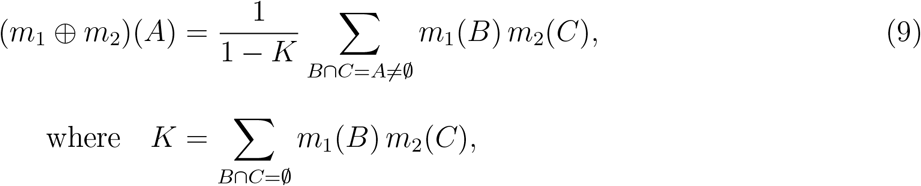

where ⊕ denotes the combination operator. Here, *m*_1_ and *m*_2_ are mass functions defined over 2^*X*^, and *A, B*, and *C* are subsets of the hypothesis space *X*. The summation is taken over all pairs of subsets whose intersection equals *A*. The term *K* is the conflict coefficient, which quantifies the degree of disagreement between the two sources of evidence; larger values of *K* indicate higher inconsistency.

The combination rule is applied iteratively to fuse the mass functions of all selected classifiers, producing a final aggregated mass distribution. From this, both the belief (*Bel*) and plausibility (*Pl*) measures can be derived, yielding lower and upper bounds on the confidence associated with each class. The final combined mass yields a single, robust estimate of the likelihood of AFib versus normal rhythm. In a nutshell, DST is an effective mechanism for integrating classifier outputs, reinforcing consistent evidence and explicitly handling uncertainty and conflict among ensemble members.

### 2.8. MOE-ECG framework for AFib detection

In this subsection, we present the workflow of the proposed MOE-ECG method for robust AFib detection. The framework utilizes multi-objective classifier selection and evidence fusion using DST. It consists of several steps, which begins with preprocessing and segmentation of ECG signals, extraction of RRI–based features, selection of an informative and non-redundant feature subset, and finally, construction of a diverse and accurate ensemble using MOPSO. The flowchart of the proposed MOE-ECG approach is illustrated in Figure 5, which is described in the following main steps:

**Figure 5:**
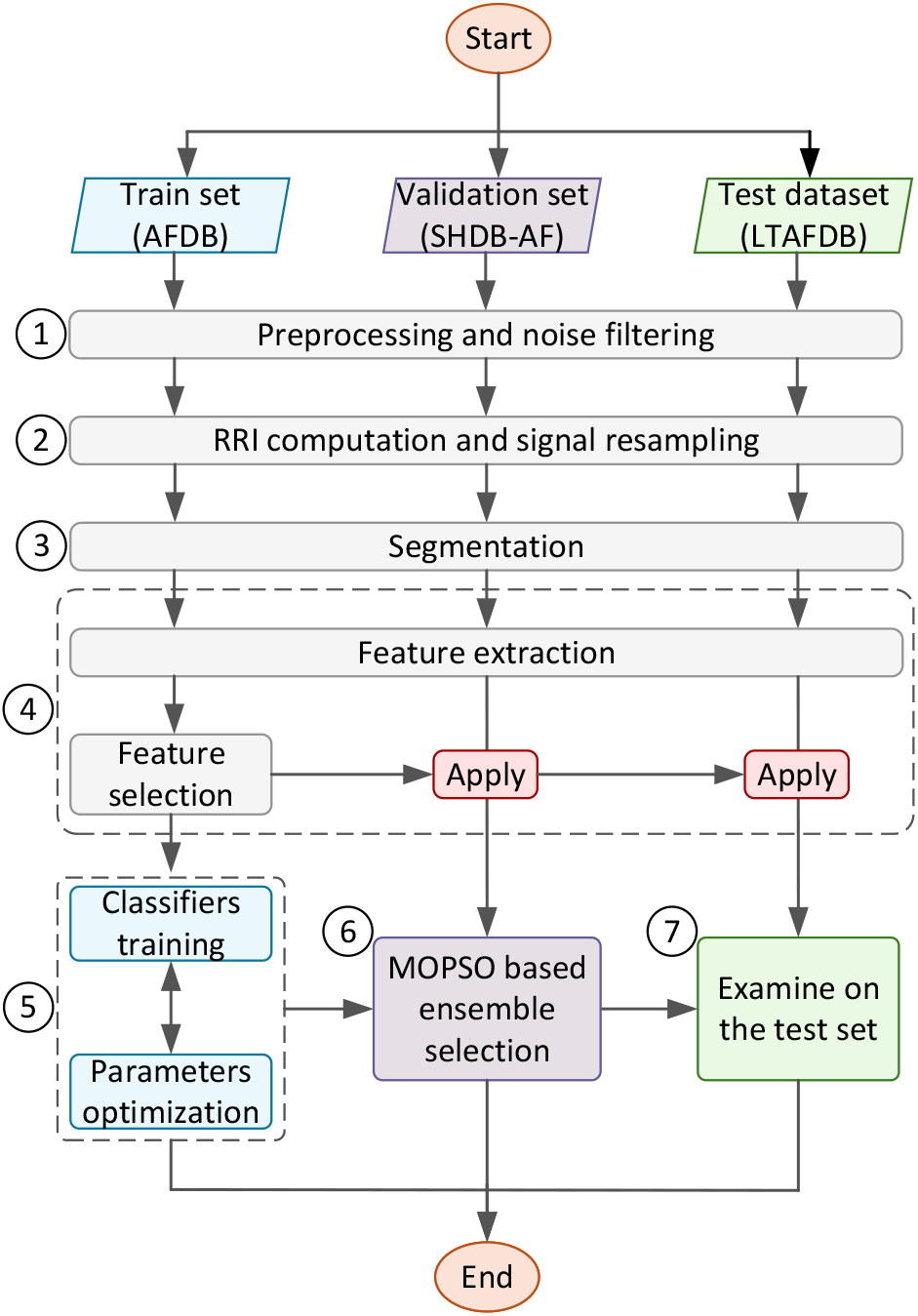
Flowchart of the proposed MOE-ECG algorithm.

1. *Preprocessing and noise filter*: All ECG recordings from the three datasets (i.e., AFDB, SHDB-AF, and LTAFDB) were filtered to remove baseline wander and high frequency noise. Then, R-peaks were extracted using the corresponding annotations files.
2. *RRI computation and signal resampling*: Each ECG recording was resampled to 250 Hz for consistency across the datasets. RRIs were computed as the time differences between consecutive R-peaks, which was used as the basis for rhythm characterization.
3. *Segmentation into 20, 60, and 120 RRIs*: The RRI sequences were segmented into windows of size 20, 60, and 120 with 50% overlap. Each segment was labeled as AFib or Normal using a majority rule threshold, where ≥ 50% AFib RRIs define an AFib window.
4. *Feature extraction and selection*: The fifteen features, as described in Subsection 2.3 and given in Table 3, were extracted from the RRIs. The most relevant features were selected using a two-stage relevance–redundancy approach, as explained in Subsection 2.3.
5. *Classifiers training and hyperparameter optimization*: A diverse pool of classifiers were trained and optimized using Grid Search and stratified 5-fold cross-validation technique on the training set (AFDB).
6. *Multi-objective ensemble classifier selection*: The MOPSO algorithm was applied to find the most accurate and diverse subset of the classifiers. All non-dominated solutions (ensembles) discovered by MOSPO were evaluated and ranked using the validation set (SHDB-AF) to select the best one.
7. *Examine on the test set*: The best solution (ensemble) found from the previous step was examined on the test set (LTAFDB) to assess its generalization performance. It should be mentioned that the prediction outputs of the single classifiers were combined using Dempster-Shafer fusion rule as described in Subsection 2.7.

## 3. Experimental Validation

As described in Section 2, the proposed MOE-ECG framework was evaluated using three well-known ECG databases for AFib detection. The AFDB [26] was employed for training the individual classifiers, and the SHDB-AF [27, 28] were used to find the best ensemble of classifiers among the non-dominated solutions. To further assess the generalizability and robustness of the resulting ensemble, an additional independent dataset, the LTAFDB [29], was used for external testing.

Various standard metrics were derived from the confusion matrix (Table 5) to evaluate model performance. Accuracy 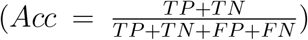, sensitivity 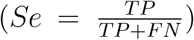, specificity 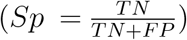, precision 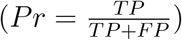, false positive rate 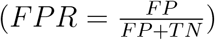, and *F*_1_-score 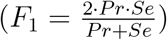 were used. Here, *TP, TN, FP*, and *FN* denote true positives, true negatives, false positives, and false negatives, respectively.

**Table 5:** Confusion matrix.

|  | Predicted negative | Predicted positive |
| --- | --- | --- |
| Actual negative | True negative (TN) | False positive (FP) |
| Actual positive | False negative (FN) | True positive (TP) |

### 3.1. Classification performance of the proposed MOE-ECG algorithm

As illustrated in Figure 6, the MOPSO algorithm produced a well-structured Pareto front, which consists of multiple non-dominated solutions shown in blue circles. Each of the non-dominated solutions represents a distinct balance between classifier performance (*F*_1_-score) and diversity. Then, all non-dominated candidate ensembles were evaluated on the validation dataset (SHDB-AF) to examine their generalization capability beyond the training set. The best-performing non-dominated solution, which is highlighted by the red star in Figure 6, was selected as the final MOE-ECG ensemble. This ensemble is used to present the performance evaluation on unseen test data (LTAFDB) using 60-beat ECG segments.

**Figure 6:**
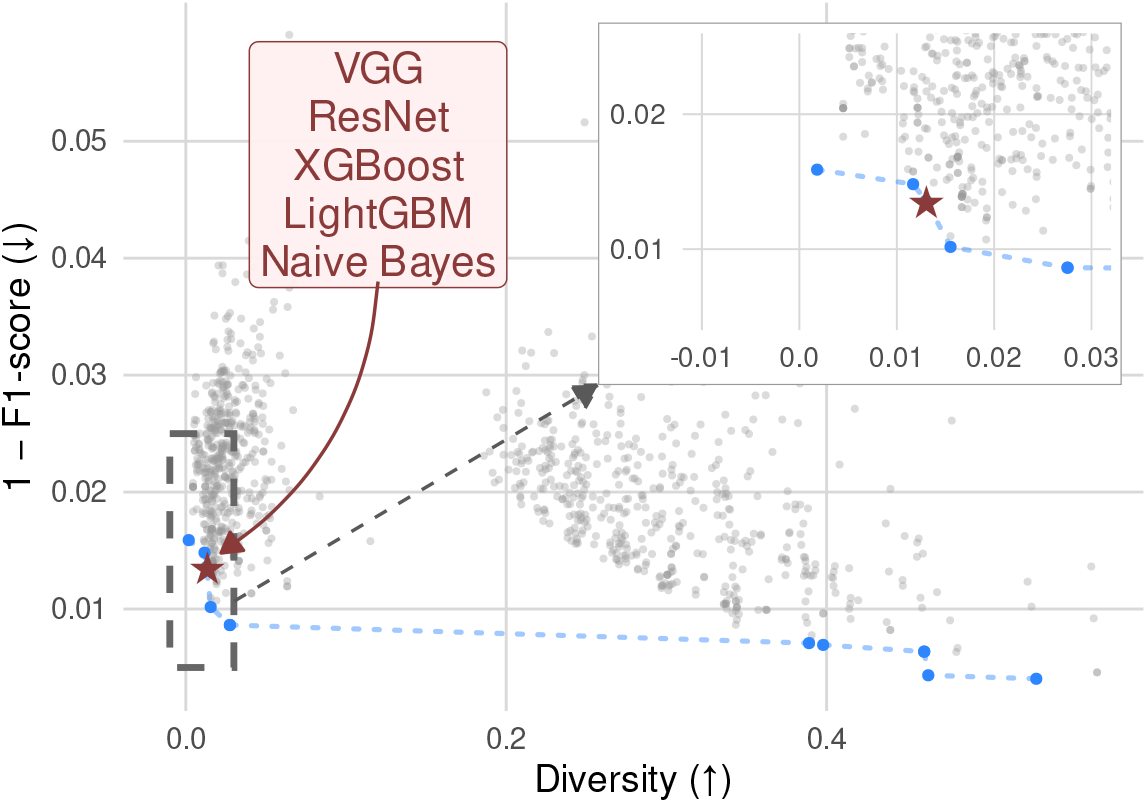
Pareto front of the MOPSO-based ensemble approach. Blue points denote non-dominated solutions, while the red star marks the selected ensemble achieving the best validation performance on SHDB-AF and used for testing on the test data (LTAFDB). The x-axis represents classifier diversity, where higher diversity indicates a more complementary ensemble, and the y-axis shows 1-F_1_-score, where lower values correspond to better classification performance.

To evaluate the performance of the proposed MOE-ECG ensemble in classifying short-term AFib, we tested its performance with input segments composed of 60 consecutive RRIs, which corresponds to a clinically actionable monitoring window. This duration is sufficient to capture irregular ventricular response patterns characteristic of AFib while remaining short enough for near-real-time screening in wearable, ambulatory, or bedside settings. Such short-term detection supports early warning, prompt confirmatory assessment, and timely initiation of preventive strategies. The final ensemble from the MOPSO based classifier selection process was evaluated on the independent LTAFDB test set to measure its capability to distinguish between AFib and Normal rhythms.

To better assess the stability of the MOPSO-based selection mechanism, Figure 7 illustrates the top-10 most frequently selected classifiers for each segment length (20, 60, and 120 RRIs). The figure highlights the dominant contributors (classifiers) to the Pareto optimal ensembles (blue circles in Figure 6) across five independent optimization runs.

**Figure 7:**
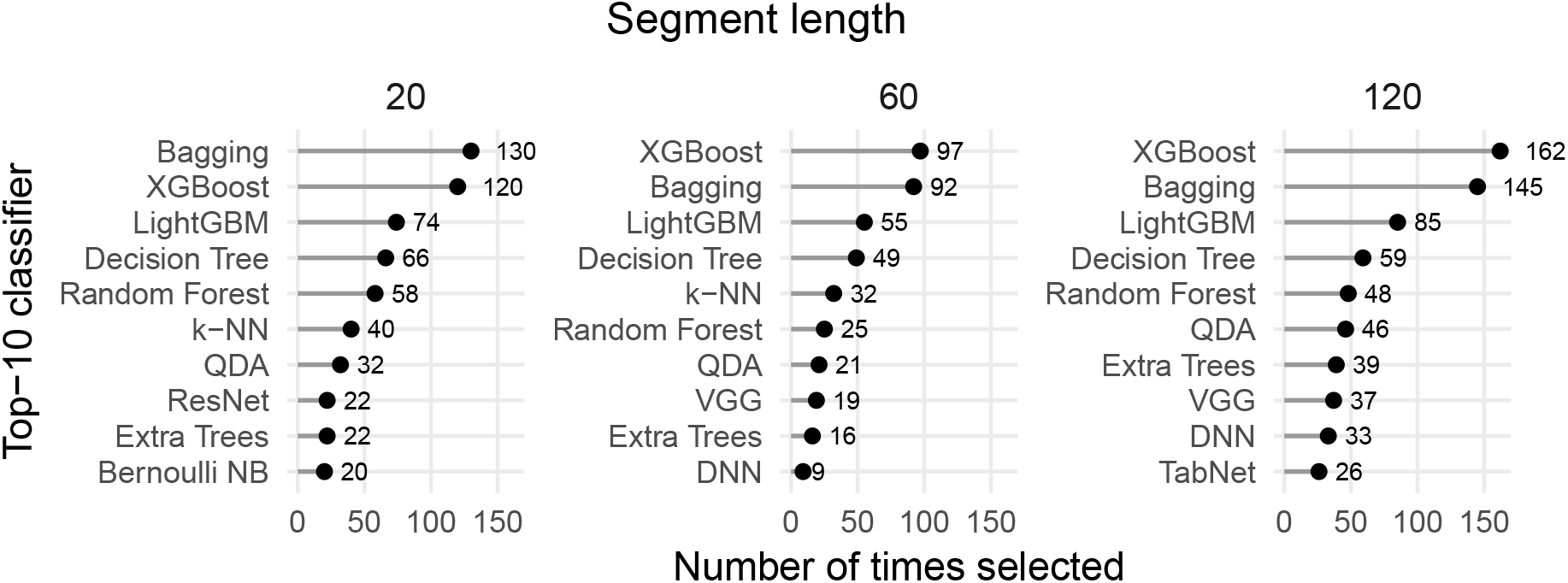
Top-10 most frequently selected classifiers across five MOPSO optimization runs for segment lengths of 20, 60, and 120 RRIs. The lollipop plot highlights the dominant contributors to the Pareto optimal solutions (blue circles in Figure 6).

Boosting and tree-based methods such as XGBoost, Bagging, LightGBM, Decision Tree, and Random Forest, consistently rank among the most frequently selected classifiers across all segment lengths. This indicates that the proposed multi-objective ensemble model favors models that achieve strong *F*_1_-score while maintaining structural diversity. On the other hand, these classifiers can efficiently capture the nonlinear and irregular characteristics of AFib cases.

Models such as QDA and k-NN appear intermittently, which suggests that complementary decision boundaries also contribute to the performance–diversity balance. In contrast, linear and naïve Bayes classifiers are rarely selected, which indicates their limited contribution under the proposed optimization criteria. Overall, Figure 7 confirms that MOPSO consistently converges toward a compact and reproducible subset of high-performing and complementary classifiers, which shows the stability of the proposed ensemble strategy.

Figure 8a presents the ROC curve of the resulting ensemble, which exhibits excellent discrimination across a wide range of decision thresholds. The curve achieves an area under the curve (AUC) of well above 0.94, which indicates the probability outputs of the model provide strong separation between AFib and NSR. As given in the figure, several operating points, which correspond to thresholds of 0.7, 0.5, and 0.2, are highlighted on the ROC curve to illustrate how different cutoff values influence sensitivity and specificity. The pink shaded regions surrounding these points represent uncertainty bands derived from bootstrapped resampling. Narrow regions indicate stable performance, whereas wider bands suggest greater sensitivity to data fluctuations. Together, these visual cues provide additional context for interpreting the Youden J point (0.8071), which corresponds to the threshold that maximizes the combined sensitivity and specificity and serves as an optimal operating point for AFib detection.

**Figure 8:**
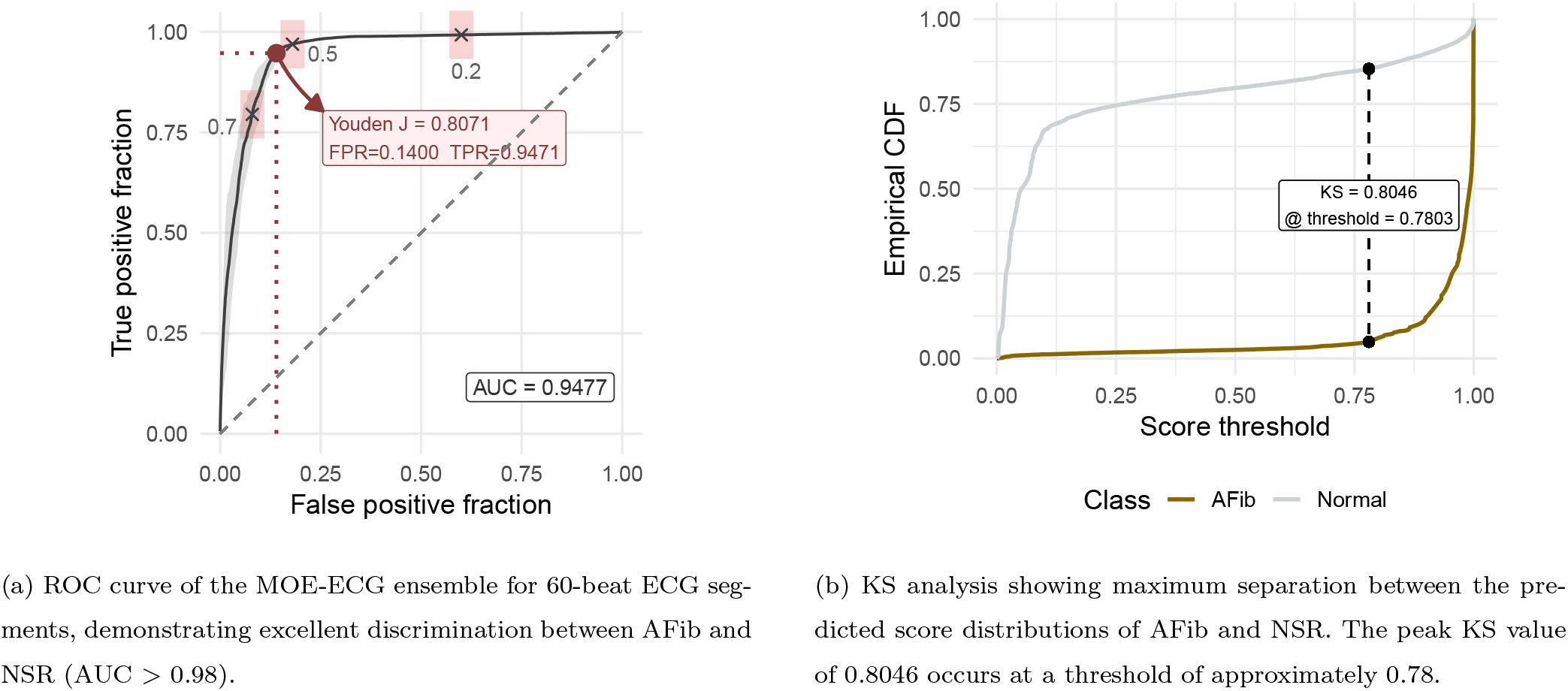
Performance evaluation of the MOE-ECG ensemble on 60-beat ECG segments. (a) ROC curve quantifying overall discrimination performance. (b) KS statistic identifying the optimal threshold that maximizes separation between AFib and NSR probability distributions. Both plots use harmonized axis scaling and grid style for visual consistency.

In addition, to derive an operationally optimal threshold value, we computed the Kolmogorov-Smirnov (KS) statistic [58], that indicates the maximum separation of empirical cumulative distribution function (CDF) [59] of AFib and Normal prediction scores. In Figure 8b, the maximum KS value of 0.8046 is obtained at the threshold of 0.7803. This large KS distance confirms that the two score distributions are strongly separated and provides a principled threshold choice for binary classification.

The optimal decision threshold obtained from the ROC analysis (Figure 8a) was determined using the Youden’s J statistic, which is defined as *J* = Se + Sp − 1 [60]. As shown in Figure 8a, the Youden’s J point occurred at a threshold equal to 0.8071, which represents the point that maximizes the balance between *Se* and *Sp*. This value is highly consistent with the threshold obtained from the KS analysis (Figure 8b), which identifies the maximum class-separation at threshold of approximately 0.78. The close agreement between these two independently derived thresholds indicates that the ensemble classifier shows stable and robust separation between AFib and Normal rhythms, with minimal sensitivity to small shifts in the decision boundary.

To further explore the effect of thresholding, three versions of the confusion matrix plot were generated, each showing how the false prediction rate varies across possible score cutoffs/thresholds. Figure 9 presents the confusion matrices for the three different thresholds of 0.5 (default), 0.8071 (Youden’s J; Figure 8a), and 0.7803 (KS analysis; Figure 8b), respectively. These figures clearly demonstrate that thresholds near 0.78-0.81 provide the best trade-off between reducing false AFib detections and preserving AFib sensitivity.

**Figure 9:**
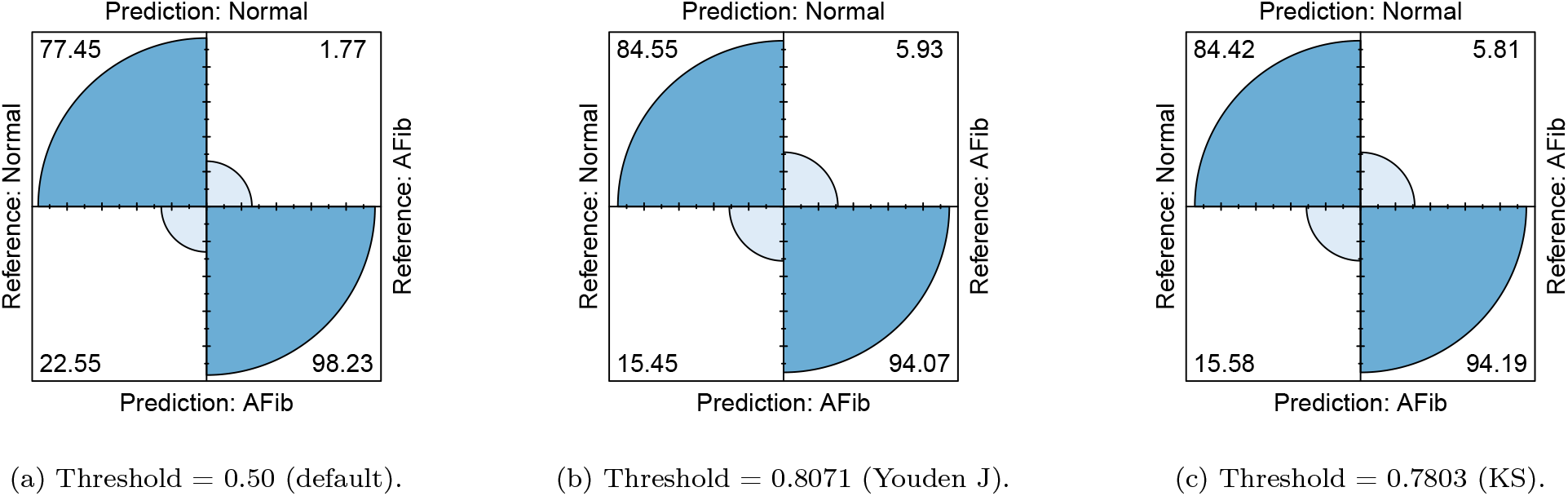
Confusion matrices of the MOE-ECG ensemble under three decision thresholds for AFib classification on 60-beat ECG segments. (a) Default threshold of 0.50. (b) Youden J optimal threshold of 0.8071, which maximizes the sum of sensitivity and specificity. (c) KS optimal threshold of 0.7803, which maximizes the separation between the predicted-score distributions of AFib and Normal rhythms. These comparisons highlight how threshold choice influences FP, FN, and overall classification performance.

Overall, the performance analysis on 60-beat ECG segments shows that the proposed MOE-ECG ensemble achieves strong separability, high stability across thresholds, and excellent classification accuracy, making it suitable for real-time AFib screening in wearable or telemonitoring applications.

### 3.2. Comparison of different segment lengths

To investigate how the temporal context of the ECG affects AFib detection, the MOE-ECG framework was evaluated using three segment lengths: 20, 60, and 120 heartbeats. Table 6 summarizes the performance of the proposed MOE-ECG model on the test set across the defined metrics. The metrics are reported as mean ± standard deviation (SD) together with the 95% confidence intervals.

**Table 6:** Comparison of MOE-ECG performance across three segment lengths on the test set (LTAFDB). Values are reported as mean (with SD and 95% CI in smaller font). Bold values indicate the best performance achieved across segment lengths for each metric.

| Metric | 20 beats | 60 beats | 120 beats |
| --- | --- | --- | --- |
| *Acc* | 0.825±0.036 [0.781, 0.870] | **0.847**±0.005 [0.841, 0.853] | 0.843±0.008 [0.833, 0.853] |
| *Se* | 0.951±0.012 [0.939, 0.967] | **0.982**±0.002 [0.979, 0.985] | 0.979±0.003 [0.975, 0.983] |
| *Sp* | 0.756±0.049 [0.695, 0.817] | **0.776**±0.008 [0.765, 0.786] | 0.765±0.014 [0.748, 0.783] |
| *Pr* | 0.684±0.044 [0.629, 0.738] | 0.697±0.008 [0.688, 0.706] | **0.705**±0.013 [0.689, 0.721] |
| *F<sub>1</sub>-score* | 0.795±0.034 [0.753, 0.838] | 0.815±0.004 [0.810, 0.821] | **0.820**±0.008 [0.810, 0.829] |
| *FPR* | 0.244±0.049 [0.183, 0.305] | **0.224**±0.008 [0.214, 0.235] | 0.235±0.014 [0.217, 0.252] |

Overall, the results show a consistent performance improvement when increasing the segment length from 20 to 60 beats, followed by a performance plateau between 60 and 120 beats. From Table 6, the 60 beats segment length gives the most balanced performance across all metrics. For example, accuracy increases from 0.825 to 0.847 when moving from 20 to 60 beats and the *F*_1_-score improves by more than 1.5%, which reflects more reliable classification between AFib and normal rhythms. Sensitivity shows a notable improvement as well, which increases from 0.951 at 20 beats to 0.982 at 60 beats.

On the other hand, increasing the window size to 120 beats does not lead to substantial improvement. For instance, accuracy, *Se*, and *Sp* remain nearly identical to those obtained with 60 beats, which confirms the hypothesis of performance saturation once sufficient temporal variability is captured. Precision shows only a marginal increase from 60 to 120 beats (i.e., from 0.697 to 0.705), which is consistent with the stable *FPR* across the longer segment lengths.

These findings indicate that 20 beat segment length, while useful for real-time detection, provides limited temporal context, which results in higher variance and reduced specificity. In contrast, 60 beat windows represent an optimal trade-off between detection performance and latency and offer the most reliable classification without the diminishing effect seen at 120 beats. Consequently, the 60 beat segment length can be selected as the primary operating point in real-world clinical settings. However, this improved performance comes at the cost of increased detection latency, as longer segments require more time to accumulate sufficient RR intervals before making a prediction. This may reduce responsiveness to rapid rhythm changes and delay early detection in time-critical scenarios, particularly in wearable or continuous monitoring settings. Consequently, while the 60 beat segment length is well-suited as a primary operating point for robust AFib detection, shorter segments such as 20 beats remain advantageous in applications requiring faster, near-real-time decision-making.

### 3.3. Comparison with the average ensemble and the single best classifier

Figure 10 presents a comparative analysis of the proposed MOE-ECG ensemble with two baseline approaches on the 60 beat ECG segments: (i) the average ensemble, which is obtained by computing the mean output across all individual classifiers, and (ii) the single best classifier, which is identified as the model with the highest *F*_1_-score on the validation data and subsequently evaluated on the unseen LTAFDB test set. The figure reports the mean classification performance across five independent runs for four key metrics (i.e., *Acc, Pr, Se*, and *F*_1_-score) for each of the three approaches.

**Figure 10:**
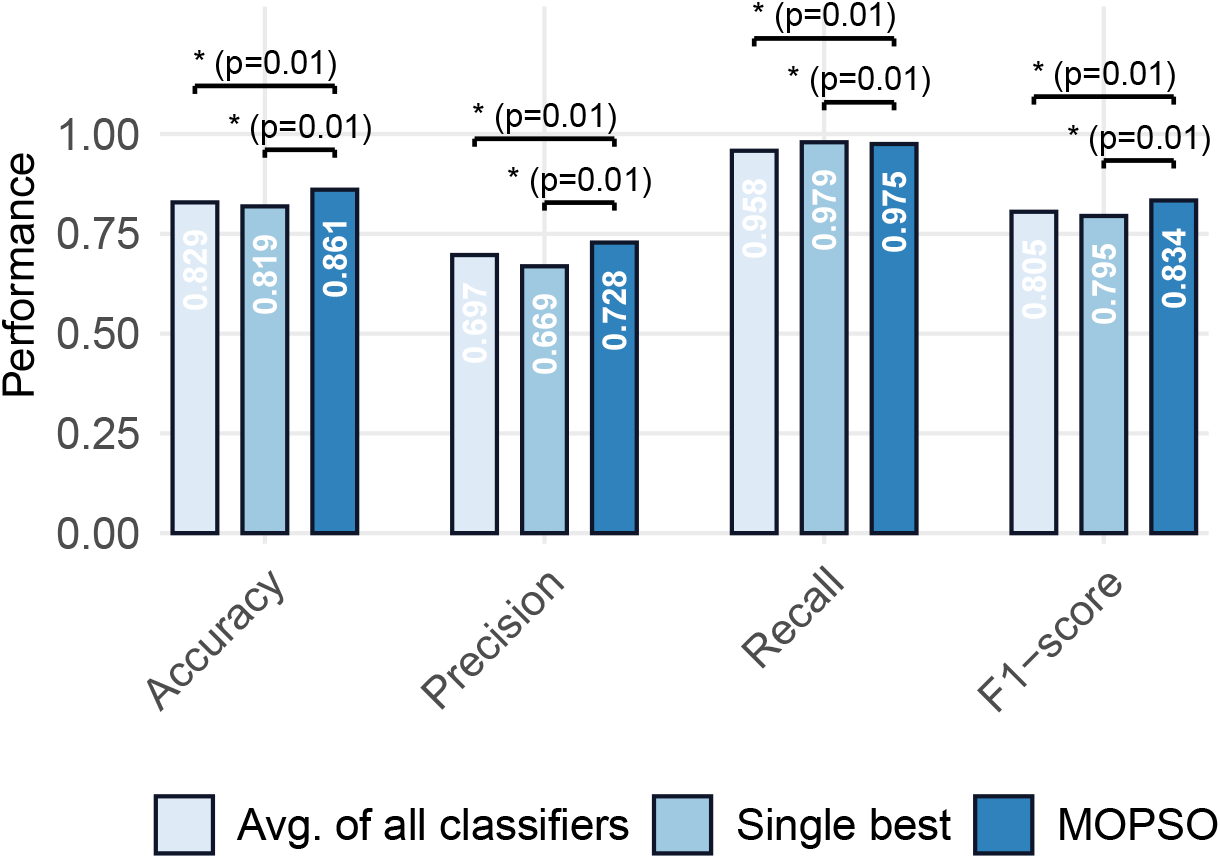
Performance comparison between the proposed MOE-ECG ensemble, the average of all classifiers, and the single best classifier on 60 beats ECG segments over four different performance metrics. The stars and connecting horizontal lines indicate the results of pairwise statistical comparisons between the three methods. Each horizontal line connects two bars whose performance distributions were compared using the Wilcoxon signed-rank test. The corresponding adjusted *p*-value (Holm correction) is displayed above the line. A single star (⋆) denotes statistical significance at the *α* = 0.05 level, which indicates that the difference in performance between the two connected methods is statistically meaningful.

Across all the metrics in Figure 10, the MOE-ECG ensemble consistently outperforms both baseline methods. The proposed MOE-ECG algorithm has performed particularly well in terms of *F*_1_-score, which confirms that MOE-ECG achieves a more favorable balance between sensitivity and precision. In other words, compared with the two baseline models, the MOE-ECG model has improved the detection of AFib episodes without compromising on the robustness to false positives. Precision also shows a substantial improvement relative to the single best classifier, which indicates that the multi-objective selection procedure and DST fusion strategy collectively reduce misclassification of normal rhythms. Although the single best model achieves marginally higher recall, this advantage is offset by its weaker precision and lower overall *F*_1_-score. The average ensemble of all classifiers generally performs worse than MOE-ECG across all metrics, which underscores the limitation of naïve averaging compared to the proposed classifier selection and fusion strategy.

To support these findings, statistical significance was applied using the Wilcoxon signed-rank test [61], which is a non-parametric paired test commonly recommended for classifier comparisons due to its robustness and distribution-free assumptions [62]. Because two pairwise comparisons were conducted (MOE-ECG vs. average ensemble and MOE-ECG vs. single best classifier), Holm’s sequentially rejective procedure [63] was applied to adjust the resulting *p*-values. All Holm-adjusted *p*-values remained below the 0.05 significance threshold, which show that the performance improvements achieved by MOE-ECG are statistically significant across repeated runs.

Overall, this comparative evaluation shows that MOE-ECG provides a substantially more accurate and reliable AFib detection system than both the average ensemble of all classifiers and the best individual classifier. These results highlight the importance of optimizing classifier diversity and evidence fusion using DST in achieving high performance and robust ECG based arrhythmia detection.

### 3.4. Performance comparison with state-of-the-art

Table 7 provides a detailed comparison of the proposed MOE-ECG model against various state-of-the-art AFib detection methods, which are all evaluated on the LTAFDB test set. The studies listed in the table represent diverse modeling approaches, which includes LSTM based recurrent neural networks, CNN and hybrid CNN-Transformer, residual and Transformer architectures, Sobolev-based learning models, and classical ensemble approaches such as AdaBoost. Although these methods differ substantially in model design, nearly all of them train on AFDB, CPSC2021 [64], or related datasets and are subsequently tested on LTAFDB, which provides a consistent benchmark for evaluating model generalization across different clinical recording environments.

**Table 7:**
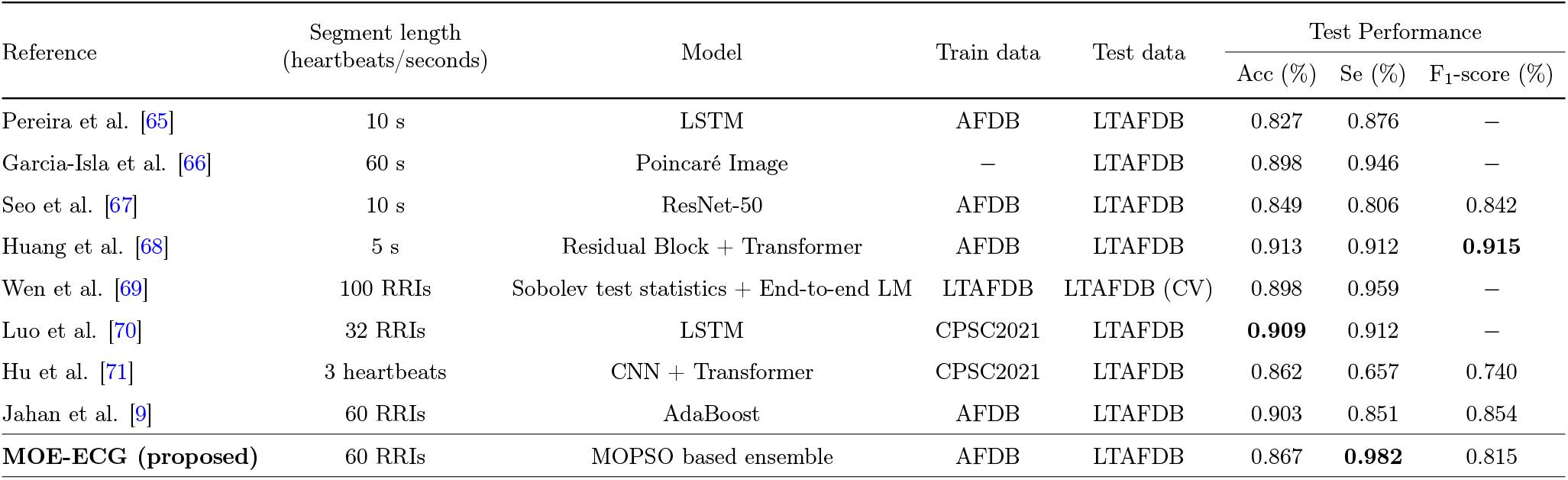
Performance Comparison of the proposed MOE-ECG model and state-of-the-art methods for AFib detection on test results. The highest values are highlighted in bold. (LSTM: Long Short-Term Memory, BiLSTM: Bidirectional LSTM, RNN: Recurrent Neural Network, CNN: Convolutional Neural Network, AdaBoost: Adaptive Boosting, LM: Learning Model, CV: Cross Validation, −: Not reported)

The reported performances show considerable variation. The accuracy, sensitivity, and *F*_1_-score values, if available, range from approximately 0.827 to 0.913, 0.657 to 0.959, and 0.740 to 0.915, respectively. As given in Table 7, models using Transformer blocks or Sobolev test-statistics tend to provide high sensitivity, while convolutional and recurrent architectures often achieve better accuracy but may be more affected by noise or domain shift when evaluated on unseen ECG recordings.

From the table, it is evident that the proposed MOE-ECG algorithm achieves highly competitive and well-balanced performance compared to existing methods. Specifically, MOE-ECG attains an accuracy, sensitivity, and *F*_1_-score of 0.867, 0.982, and 0.815, respectively. Notably, the proposed method achieves the highest sensitivity among all compared approaches, which is particularly important in AFib detection, where minimizing missed events is clinically critical. While some deep learning models report slightly higher accuracy or *F*_1_-scores, these models often rely on complex architectures such as CNNs, LSTMs, or Transformers, which require large-scale training data and high computational resources.

In contrast, the proposed MOE-ECG framework adopts a feature-based and diversity-aware ensemble strategy, which enables robust generalization across datasets while maintaining lower computational complexity. By leveraging a compact set of informative RRI-based features and selecting a small subset of complementary classifiers from an initial pool of 22 models, the framework avoids the need for deep end-to-end inference while still capturing nonlinear and heterogeneous ECG patterns. This contributes to improved robustness under domain shift, as evidenced by strong performance on the unseen LTAFDB dataset.

Furthermore, the proposed approach is inherently suitable for real-time and resource-constrained applications. The use of low-dimensional feature representations and lightweight classifiers significantly reduces inference complexity compared to deep neural network models, which typically involve millions of parameters and require GPU acceleration. In contrast, the selected ensemble in MOE-ECG operates on a small number of features and classifiers, which enables fast prediction with minimal computational overhead. Combined with the use of 60 beat segments, which provide a balance between detection accuracy and latency, this makes the proposed framework well-suited for deployment in wearable devices and continuous monitoring systems.

Overall, Table 7 demonstrates that MOE-ECG not only achieves competitive performance compared to state-of-the-art deep learning and statistical models under the same evaluation conditions, but also offers advantages in terms of robustness, interpretability, and computational efficiency. These properties make it a practical and scalable solution for real-world AFib detection.

## 4. Conclusion

This study proposes a multi-objective ensemble fusion framework (MOE-ECG), designed to enhance the generalizability and robustness of AFib detection from short ECG segments. This model combines twenty two classifiers to create an ensemble, which is simultaneously diverse and accurate, using MOPSO algorithm. The model selects subsets of base learners and combines their outputs using DST to produce reliable probabilistic decisions.

Various RRI features were extracted from three different ECG datasets: AFDB, SHDB-AF, and LTAFDB. These datasets were used for training, validation, and independent testing, respectively. The proposed MOE-ECG model showed robust performance consistently. Three different segment lengths (i.e., 20, 60, and 120 beats) were examined to exhibit the robustness of the model. The 60-beat segments were chosen as the optimal operating window, which provides the best balance between temporal context and detection latency. The selected ensemble by MOPSO algorithm achieved an accuracy, sensitivity, and *F*_1_-score of 0.867, 0.982, and 0.815, respectively, which outperforms both the best individual classifier and the average ensemble of all classifiers.

In addition, the results of the MOE-ECG model were compared with state-of-the-art AFib detection approaches, which highlight the comparable performance of the proposed algorithm. For example, it offers the highest sensitivity among all published methods, which shows its capability to minimize missed AFib episodes. The stability of thresholding results, the consistent recurrence of diverse high-performing classifiers in the Pareto-optimal solutions, and significant improvements confirmed through Holm-adjusted Wilcoxon tests collectively validate the robustness of the proposed multi-objective optimization and fusion framework.

Overall, the experimental results support that MOE-ECG is an effective and robust solution for AFib detection in short episodes of ECGs. This is particularly important for real-time monitoring in wearable and telehealth applications. For example, the optimal operating window of 60s is used in single-lead, handheld ECG devices for primary care [11], where MOE-ECG could easily be adjusted to. In addition to AFib detection, the proposed approach can be extended to other cardiac arrhythmias diagnosis, which requires robustness across devices and populations.

There are also some limitations for the study, which can be addressed in future studies. First, in this study, only RRI-based features were used that can be extended by including other types of features. Second, more datasets can be added for testing the proposed model to better present its generalizability on diverse populations. Future work will explore multi-lead ECGs, adaptive windowing, and on-device deployment by reducing the complexity of the model. Additionally, the proposed model can be compared with state-of-the-art ECG foundation models and other large or small language models (LLMs/SLMs) to extract relevant features directly from raw ECGs, which will be useful for personalized decision support systems in resource-constrained settings.

## Data Availability

The ECG datasets used in this study can be found at https://physionet.org/content/afdb/1.0.0/, https://physionet.org/content/shdb-af/1.0.1/, and https://physionet.org/content/ltafdb/1.0.0/.

https://physionet.org/content/shdb-af/1.0.1/

https://physionet.org/content/ltafdb/1.0.0/

